# Urinary CD4^+^ Effector Memory CD38^+^ HLA-DR^+^ T Cells for Diagnosis of Acute Interstitial Nephritis

**DOI:** 10.64898/2026.06.25.26356554

**Authors:** Wenxin Sha, Pouneh Mirkheshti, Shi Feng, Christopher M. Skopnik, Jeremy Russ, Christoph Daniel, Kerstin Amann, Joram Arzig, Nina Goerlich, Sandra M. Herrmann, Jan Klocke, Jianghua Chen, Kai-Uwe Eckardt, Hong Jiang, Philipp Enghard

**Author notes:** These authors share first and last authorship, respectively (Wenxin Sha, Pouneh Mirkheshti, Hong Jiang, Philipp Enghard). **Corresponding author** *Prof. Hong Jiang* Kidney Disease Center, the First Affiliated Hospital, College of Medicine, Zhejiang University, Qingchun Road 79, Hangzhou, 310003, China., *Prof. Philipp Enghard* Department of Nephrology and Medical Intensive Care, Charité - Universitätsmedizin Berlin Luisenstraße 13, 10117 Berlin, Germany.

## Abstract

**Introduction:** Acute interstitial nephritis is an important differential diagnosis in patients with deteriorating kidney function. Diagnosis currently requires kidney biopsy, an invasive procedure associated with risks. We hypothesized that urinary T cells may serve as a non-invasive biomarker for acute interstitial nephritis.

**Methods:** A total of 320 patients undergoing clinically indicated kidney biopsy were enrolled in a discovery cohort at Charité Berlin (n = 80), an internal validation cohort at Charité (n = 100), and an external validation cohort at The First Affiliated Hospital, Zhejiang University School of Medicine, Hangzhou (n = 140). Urinary immune cells were assessed by flow cytometry. Renal T cell infiltration was evaluated by immunofluorescence in kidney biopsy specimens from the discovery and internal validation cohorts, including 16 patients with acute interstitial nephritis and 9 patients without acute interstitial nephritis. Additionally, CXCL9 was measured by ELISA in 102 urine samples from these cohorts.

**Results:** Across all cohorts, 27 patients (8.4%) were diagnosed with acute interstitial nephritis. In the discovery cohort, multiple urinary T cell subsets were increased in acute interstitial nephritis, with activated CD4^+^ effector memory T cells expressing CD38 and HLA-DR showing the strongest diagnostic performance. This marker outperformed urinary monocytes, eosinophils, and CXCL9 and was validated in both independent cohorts. Across all cohorts, the area under the receiver operating characteristic curve was 0.84 and increased to 0.91 after exclusion of 8 patients receiving corticosteroids. A cutoff of 211 activated CD4^+^ effector memory T cells per 100 mL urine yielded a sensitivity of 78% and a specificity of 81%. Urinary activated CD4^+^ effector memory T cell counts correlated with renal CD4^+^ and CD4^+^ CD38^+^ T cell infiltration in acute interstitial nephritis. **Conclusions** Urinary activated CD4^+^ effector memory T cells expressing CD38 and HLA-DR represent a promising non-invasive biomarker for the diagnosis of acute interstitial nephritis.

**Lay Summary:** Acute interstitial nephritis is a frequent cause of acute kidney injury and is usually diagnosed by kidney biopsy, an invasive procedure that carries risks. We investigated whether immune cells in urine could help identify AIN in a less invasive manner. In this multicenter study, urine samples from 320 patients undergoing kidney biopsy in Germany and China were analyzed. We found that a specific population of activated T cells was markedly increased in the urine of patients with AIN compared with patients with other kidney diseases. This urinary marker outperformed other immune cell types and the inflammatory cytokine CXCL9 and was successfully validated in independent cohorts from both Germany and China. Importantly, urinary activated T cell counts correlated with immune cell infiltration within the kidney. These findings suggest that urinary activated T cells may serve as a non-invasive biomarker to support the diagnosis of AIN and facilitate earlier clinical decision-making.

## Introduction

Acute interstitial nephritis (AIN) is an inflammatory condition of the renal interstitium that can arise from various causes, including medications, infections, and autoimmune disorders.^1,2^ In most cases, withdrawal of the offending drug and corticosteroid therapy lead to favorable clinical outcomes.^2,3^ However, AIN usually presents with an elevation of serum creatinine concentrations accompanied by either no or nonspecific clinical features, such as oliguria, proteinuria and edema,^1,4^ resulting in diagnostic challenges and treatment delays. At present, confirmative diagnosis relies on kidney biopsy, an invasive procedure that carries inherent risks.^5^ As a result, despite accounting for up to 10-20% of acute kidney injury (AKI) cases, AIN remains frequently undiagnosed.^6,7^ Therefore, there is an urgent clinical need for rapid and non-invasive biomarkers to facilitate the diagnosis and therapeutic monitoring of AIN.

In previous studies, eosinophils detected in urine sediment were reported as a potential diagnostic marker for AIN.^8^ However, a retrospective analysis by Muriithi et al. reported a sensitivity of 30.8 % and a specificity of 68.2 %, indicating the inadequacy of this semiquantitative methodology.^9^ In consideration of the inflammatory nature of AIN, which is characterized by cytokine-mediated immune activation, attention has increasingly shifted toward chemokines as potential diagnostic biomarkers.^10^ Recent studies by Moledina et al. have reported urinary IL-9, TNF-α, and CXCL9 as promising candidates.^11,12^ In particular, urinary CXCL9 has been reported and validated as a diagnostic biomarker for AIN across several studies.^12–15^

Given the fact that the histopathological hallmark of AIN is an interstitial lymphocytic infiltrate,^1^ and our previous findings suggesting that urinary T cells reflect renal inflammation in various inflammatory kidney diseases^16–18^, we hypothesized that urinary immune cell counts may serve as a biomarker of ongoing renal interstitial inflammation. Here, we report the results of a multicenter prospective AIN biomarker study evaluating the diagnostic value of urinary T cells in general, CD4^+^ effector memory (EM) CD38^+^ HLA-DR^+^ T cells, monocytes and eosinophils, as potential biomarkers for AIN. We compared our results with urinary CXCL9 concentrations.

## Methods

### Study design and participants

This AIN biomarker study is a multicenter study consisting of a discovery cohort and an internal validation cohort from Charité University Medical Center in Berlin, Germany (Campus Berlin Mitte and Campus Virchow Klinikum), as well as an external validation cohort from The First Affiliated Hospital, Zhejiang University School of Medicine (FAHZU), Hangzhou, China (**Supplementary Figure S1**). The study was registered in January 2025 (DRKS00035356). Ethical approval was obtained at all participating centers (EA4/203/24; 2024-1420), and all participants or their legal guardians provided informed consent.

Inclusion criterion was a clinically indicated kidney biopsy in individuals with an acute, unexplained reduction in glomerular filtration rate. Exclusion criteria were age < 18 years at Charité or < 14 years at FAHZU, kidney transplantation, urinary tract infection, menstruation, cancellation of kidney biopsy after urine collection, insufficient biopsy tissue, or inconclusive histopathological diagnosis (**Supplementary Figure S1**).

Between May 2023 and June 2024, 91 patients were recruited into the discovery cohort, and between June 2024 and July 2025, 124 patients were enrolled into the internal validation cohort. Between October 2024 and April 2025, 159 patients were enrolled into the external validation cohort. The target sample size for the internal validation cohort was determined a priori in consultation with a biostatistician from the Institute of Biometry and Clinical Epidemiology (iBikE), Charité, based on the diagnostic performance observed in the discovery cohort. After application of predefined exclusion criteria, 80, 100, and 140 patients from the discovery, internal validation, and external validation cohorts, respectively, were included in the final analysis (**Supplementary Figure S1**).

### Stratification of patients by diagnostic groups

Diagnoses were assigned according to kidney biopsy reports. Patients were classified into three groups: (1) biopsy-confirmed AIN without primary glomerular disease, (2) glomerular disease with concomitant lymphocytic interstitial infiltrate (LI), and (3) kidney diseases without evidence of interstitial inflammation (OD) (**Supplementary Table S1**).

### Urine sample collection and processing

Urine samples were collected within 72 hours before kidney biopsy whenever possible and transported at 4 °C. If pre-biopsy samples were unavailable, urine was collected between 24 hours and 7 days after biopsy. Eight samples were obtained after initiation of corticosteroid therapy. Samples were processed using a previously validated preservation protocol for urinary immune cells^19^ and subsequently stored at -80 °C until analysis. Detailed information on sample preservation, storage conditions, and buffer composition is provided in the **Supplementary Methods**. Unfixed urinary supernatants intended for CXCL9 measurements were aliquoted and stored at -80 °C.

### Urinary immune cell staining and flow cytometry

Frozen urine samples were thawed, filtered through a 30 µm cell strainer, centrifuged, and resuspended in phosphate-buffered saline containing bovine serum albumin and ethylenediaminetetraacetic acid. In the discovery cohort, monocyte/eosinophil and T cell staining panels were performed, whereas only the T cell panel was applied in the internal and external validation cohorts. Following Fc receptor blockade, samples were stained with fluorochrome-conjugated antibodies and analyzed using a BD LSRFortessa Cell Analyzer (Berlin) or Cytek Aurora Analyzer (FAHZU). Data were analyzed using FlowJo software (version 10.8). Antibodies, staining conditions, panel composition, and gating strategies are summarized in **Figure 1a, Supplementary Table S2** and **Supplementary Figure S2**. Immune cell counts were normalized to 100 mL urine by accounting for urine volume and processed sample fractions.

**Figure 1.**
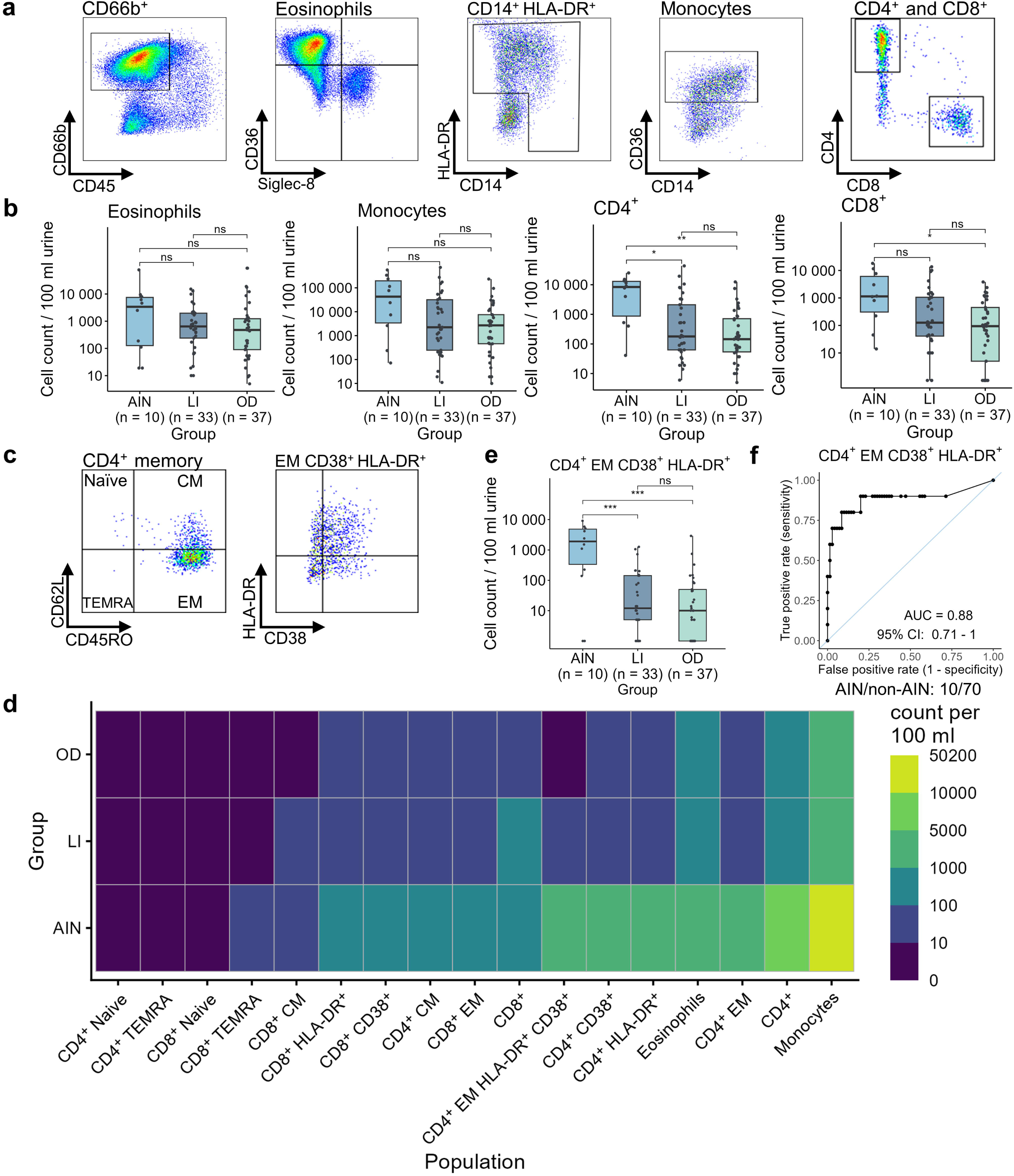
Identification of urinary CD4^+^ EM CD38^+^ HLA-DR^+^ T cells as a potential biomarker for AIN in the discovery cohort. (a) Gating strategies for urinary eosinophils, monocytes, CD4^+^, and CD8^+^ T cells. (b) Comparison of eosinophils, monocytes, and T cell counts per 100 mL urine between diagnosis groups. (c) Identification of CD4^+^ EM CD38^+^ HLA-DR T cells as the best T cell subset to discriminate AIN from LI and OD. (d) Heatmap shows abundance of immune cell subsets per 100 mL urine in different diagnosis groups. (e) Comparison of corresponding counts per 100 mL urine between diagnosis groups. (f) ROC curve analysis evaluates the diagnostic performance of urinary CD4^+^ EM CD38^+^ HLA-DR T cells in predicting AIN. Significance levels: ns, no significance, \**P* < 0.05, \*\**P* < 0.01, \*\*\**P* < 0.001. AIN, acute interstitial nephritis; LI, lymphocytic interstitial infiltrate in primary glomerular diseases; OD, other diseases without interstitial involvement; CM, T central memory cells; EM, T effector memory cells; TEMRA, T effector memory cells re-expressing CD45RA; ROC, receiver operating characteristic; AUC, area under the curve; CI, confidence interval.

### Immunofluorescence staining of renal T cells

Immunofluorescence staining for CD4 and CD38 was performed on kidney biopsy specimens from 16 patients with AIN and 9 patients without AIN from the discovery and internal validation cohorts. Digital image acquisition and quantification were performed using QuPath software.^20^ Detailed staining procedures, antibodies, image acquisition, and image analysis methods are described in the **Supplementary Methods**.

### Validation of urinary CXCL9 as potential AIN biomarker

Urinary CXCL9 concentrations were measured in 45 patients from the discovery cohort and 57 patients from the internal validation cohort using a commercially available immunoassay according to the manufacturer’s instructions. Urinary creatinine concentrations were measured to normalize CXCL9 levels. Detailed assay methodology, validation experiments, and data processing procedures are described in the **Supplementary Methods and Supplementary Figure S3.**

### Clinical data collection

Demographic information (age and gender), laboratory parameters of kidney function, medication history, diagnostic information in kidney biopsy reports, and digitized images of renal biopsy slides were collected from the electronic medical record systems at Charité Berlin and FAHZU, as well as the respective departments of pathology. Laboratory indices included serum creatinine (sCr), estimated glomerular filtration rate (eGFR, calculated using the CKD-EPI formula 2021), protein-to-creatinine ratio (PCR), and albumin-to-creatinine ratio (ACR). In order to identify potential factors contributing to the development of AIN or affecting our experimental results, medication history within 60 days prior to current admission was additionally collected for the AIN group.

### Statistical analysis

All statistical analyses were conducted using R software (version 4.3.3) and RStudio (version 2023.03.1+466). Most data transformation was performed with packages from the “tidyverse”, flow cytometric data extraction from FlowJo was performed using the “fcexpr” package, and data visualization was realized with “ggplot2” and “ggpubr”. Continuous variables were presented as mean (SD) for normally distributed data and median (IQR) for skewed distributions, while categorical variables were presented as counts (percentages).

For pairwise comparisons of immune cell counts and CXCL9 levels among diagnosis groups (AIN, LI and OD), the *Wilcoxon rank-sum* test was applied, followed by *Benjamini-Hochberg*-adjusted pairwise comparisons for the discovery cohort and *Bonferroni*-adjusted pairwise comparisons for both validation cohorts and pooled analyses across all cohorts. To compare differences of multiple biomarkers among groups within the discovery cohort, a heatmap displaying group medians was generated using the “pheatmap” package, as the data exhibited skewed distributions. After identifying the optimal biomarker, receiver operating characteristic (ROC) curves and area under the curve (AUC) values with confidence intervals (CI) were generated using the “pROC” package. Within the AIN group, the *Wilcoxon rank-sum* test was used to compare biomarker levels between patients with or without corticosteroid treatment prior to sample collection. Correlations between lymphocytic infiltration grades and biomarker levels in AIN patients were assessed using *Spearman* correlation analysis based on the “psych” package. Statistical power calculations for ROC curves were performed using the “pROC” package.^21^ All tests were two-sided, and *P* values < 0.05 were considered statistically significant.

## Results

### Participant characteristics

Baseline clinical characteristics of the 320 participants from the discovery, internal validation, and external validation cohorts (**Supplementary Figure S1**) are summarized in **Table 1**. The median age was 50 years (IQR, 35–61), and 181 participants (56.6%) were men. According to kidney biopsy results, patients were classified into three groups: (1) biopsy-confirmed acute interstitial nephritis (AIN) without primary glomerular disease, (2) glomerular disease with concomitant lymphocytic interstitial infiltrate, and (3) kidney diseases without evidence of interstitial inflammation (OD). Overall, 27 patients (8.4%) had AIN, 98 (30.6%) had LI, and 195 (61.0%) were classified as OD. Detailed distributions of biopsy diagnoses are provided in **Table 1**. The discovery cohort included 10 patients with AIN, 33 with LI, and 37 with OD; the internal validation cohort 7, 45, and 48 patients, and the external validation cohort 10, 20, and 110 patients, in the three categories, respectively. IgA nephropathy was the most common primary biopsy diagnosis (89 patients, 27.8%), particularly in the external validation cohort (59 patients, 42.1%), followed by diabetic and/or hypertensive nephropathy (77 patients, 24.0%).

**Table 1.** Baseline characteristics of the AIN study cohort.

|  |  | Overall<br>N = 320 | Discovery<br>cohort<br>N = 80 | Internal<br>validation<br>cohort<br>N = 100 | External<br>validation<br>cohort<br>N = 140 |
| --- | --- | --- | --- | --- | --- |
| Age (mean [SD]),<br>(median [IQR]) |  | 48 [16],<br>50 [35-61] | 50 [17],<br>51 [37-63] | 52 [15],<br>54 [39-63] | 44 [16],<br>45 [31-57] |
| Gender (%) | Male | 181 (56.6) | 47 (58.7) | 62 (62.0) | 72 (51.4) |
|  | Female | 139 (43.4) | 33 (41.3) | 38 (38.0) | 68 (48.6) |
| Serum creatinine, mg/dL<br>(median [IQR]) |  | 1.34 [0.89-2.20] | 1.83 [1.27-3.48] | 1.67 [0.98-2.60] | 1.09 [0.78-1.53] |
| eGFR, (median [IQR]) | mL/min per<br>1.73 m<sup>2</sup> | 55 [32-93] | 42 [19-56] | 45 [23-82] | 73 [50-104] |
| PCR,<br>(median [IQR]) | mg/g creatinine | 1094 [391-3383] | 1399 [290-4130] | 1128 [458-4530] | 1020 [470-2325] |
| ACR,<br>(median [IQR]) | mg/g creatinine | 849 [200-2552] | 948 [72-3342] | 734 [150-3258] | 867 [293-1933] |
| Primary disease (%) | AIN | 27 (8.4) | 10 (12.5) | 7 (7.0) | 10 (7.1) |
|  | IgAN | 89 (27.8) | 4 (0.5) | 26 (26.0) | 59 (42.1) |
|  | Diabetic/hypertensive | 77 (24.0) | 34 (42.5) | 26 (26.0) | 17 (12.1) |
|  | Membranous GN | 27 (8.4) | 9 (11.3) | 5 (5.0) | 13 (9.2) |
|  | Minimal change | 20 (6.3) | 4 (0.5) | 7 (7.0) | 9 (6.4) |
|  | ATI | 14 (4.3) | 5 (6.2) | 6 (6.0) | 3 (2.1) |
|  | Lupus Nephritis | 6 (1.9) | 0 (0.0) | 6 (6.0) | 0 (0.0) |
|  | Amyloidosis | 5 (1.6) | 0 (0.0) | 2 (2.0) | 3 (2.1) |
|  | ANCA | 2 (0.6) | 0 (0.0) | 1 (1.0) | 1 (0.7) |
|  | Other<sup>a</sup> | 53 (16.6) | 14 (17.5) | 14 (14.0) | 25 (17.9) |
| Group (%) | AIN | 27 (8.4) | 10 (12.5) | 7 (7.0) | 10 (7.1) |
|  | LI | 98 (30.6) | 33 (41.2) | 45 (45.0) | 20 (14.3) |
|  | OD | 195 (61.0) | 37 (46.3) | 48 (48.0) | 110 (78.6) |

<sup>a</sup>Other primary kidney diseases include, but are not limited to Henoch-Schonlein purpura, focal segmental glomerulosclerosis, light chain deposition disease, and obesity-related glomerulopathy. sCr, serum creatinine; eGFR, estimated glomerular filtration rate (using the CKD-EPI-formula); PCR, protein-to-creatinine ratio; ACR, albumin-to-creatinine ratio; ATI, acute tubular injury; AIN, acute interstitial nephritis; ANCA, ANCA-associated vasculitis; IgAN, IgA nephropathy; GN, glomerulonephritis; LI, patients with lymphocytic interstitial infiltrates in a primary glomerular disease; OD, patients with other kidney diseases without interstitial involvement.

### Urinary CD4^+^ EM CD38^+^ HLA-DR^+^ T cells are the best subset to differentiate AIN from other entities

Urinary monocyte and eosinophil counts did not show significant differences among the AIN, LI, and OD groups (**Figure 1b**). In contrast, urinary CD4^+^ and CD8^+^ T cell counts per 100 mL urine were significantly increased in patients with AIN compared with those in OD, and urinary CD4^+^ T cell counts also distinguished AIN from LI (**Figure 1b**). To further identify T cell subsets with higher discriminatory potential for AIN, urinary T cells were classified into naïve, central memory (CM), effector memory (EM), and terminally differentiated effector memory cells re-expressing CD45RA (TEMRA) based on CD45RO and CD62L expression. EM T cells were then further stratified according to CD38 and HLA-DR expression as markers of activation (Figure 1C). Median counts of different immune cell populations across diagnostic groups are summarized in a heatmap (**Figure 1d**). Among these subsets, urinary CD4^+^ EM CD38^+^ HLA-DR^+^ T cells emerged as the best T cell subset to discriminate AIN from LI and OD (*P* < 0.001; **Figure 1e**). Representative Hematoxylin- and Eosin-stained kidney biopsy sections from patients with AIN, LI, and OD, together with corresponding flow-cytometric profiles after gating of CD4^+^ EM T cells, are shown in **Supplementary Figure S4**. In the discovery cohort, urinary CD4^+^ EM CD38^+^ HLA-DR^+^ T cells achieved an area under the receiver operating characteristic curve (ROC-AUC) of 0.88 (95% CI, 0.71–1.00; **Figure 1f**).

### Internal and external validation of urinary CD4^+^ EM CD38^+^ HLA-DR^+^ T cells as a potential AIN biomarker

The diagnostic performance of urinary CD4^+^ EM CD38^+^ HLA-DR^+^ T cells was independently validated in both the internal and external validation cohorts (**Figure 2a and c**), with ROC-AUCs of 0.77 (95% CI, 0.54-0.99) and 0.85 (95% CI, 0.71-0.99), respectively (**Figure 2b and d**). In a pooled analysis of the discovery and both validation cohorts, urinary CD4^+^ EM CD38^+^ HLA-DR^+^ T cells retained good discriminatory performance for AIN (AIN vs. OD: *P* < 0.0001 and AIN vs. LI: *P* < 0.0001; **Figure 2e**), yielding an overall ROC-AUC of 0.84 (95% CI, 0.74-0.93; **Figure 2f**). Using a cutoff value of 211 activated CD4^+^ EM T cells per 100 mL urine, as determined by Youden’s index, resulted in a sensitivity of 78% and a specificity of 81%.

**Figure 2.**
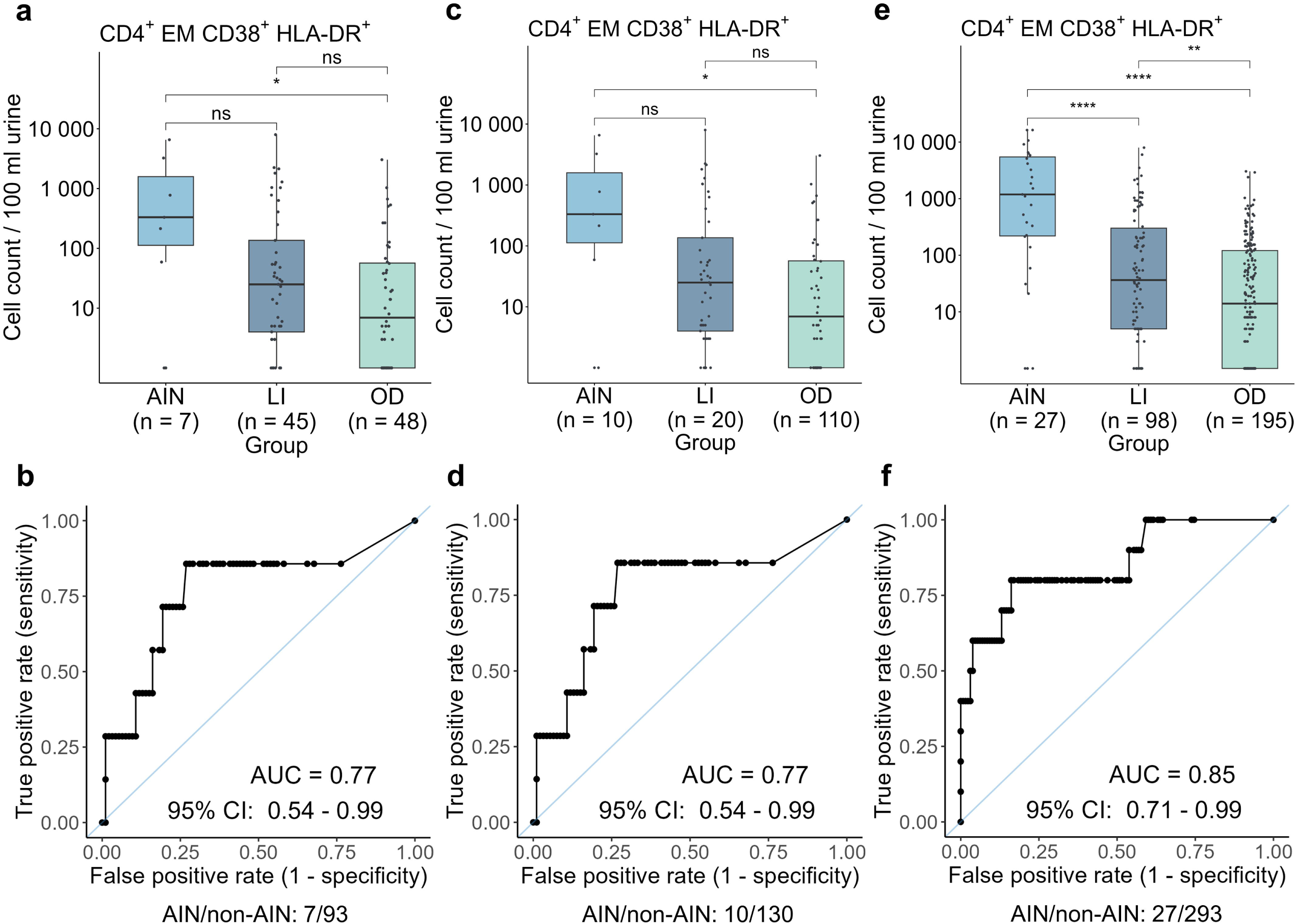
Internal and external validation of urinary CD4^+^ EM CD38^+^ HLA-DR^+^ T cells as a potential biomarker for AIN. Comparison of urinary CD4^+^ EM CD38^+^ HLA-DR T cell counts per 100 mL of urine between diagnosis groups and ROC curve analysis showing the diagnostic performance of this subset in predicting AIN in the (a, b) internal validation cohort, (c, d) external validation cohort, and (e, f) all three cohorts. Significance levels: ns, no significance, \**P* < 0.05, \*\**P* < 0.01, \*\*\**P* < 0.001, \*\*\*\**P* < 0.0001. AIN, acute interstitial nephritis; LI, lymphocytic interstitial infiltrate in primary glomerular diseases; OD, other diseases without interstitial involvement; ROC, receiver operating characteristic; AUC, area under the curve; CI, confidence interval.

### Urinary CXCL9 fails to distinguish AIN from other kidney diseases

Since previous reports suggested urinary CXCL9 as a potential biomarker for AIN, we additionally evaluated its diagnostic performance in our cohorts. Urinary CXCL9 levels were measured in 45 samples from the discovery cohort and 57 samples from the internal validation cohort. In contrast to urinary CD4^+^ EM CD38^+^ HLA-DR^+^ T cells, normalized urinary CXCL9 levels (pg / mg creatinine) did not differ significantly among the diagnostic groups (*P* > 0.1 for all comparisons; **Figure 3a**) and showed limited discriminatory performance for AIN, with a ROC-AUC of 0.68 (95% CI, 0.5-0.86; **Figure 3b**). Overall, urinary CD4^+^ EM CD38^+^ HLA-DR^+^ T cells outperformed urinary CXCL9 in discriminating AIN from other diagnostic groups.

**Figure 3.**
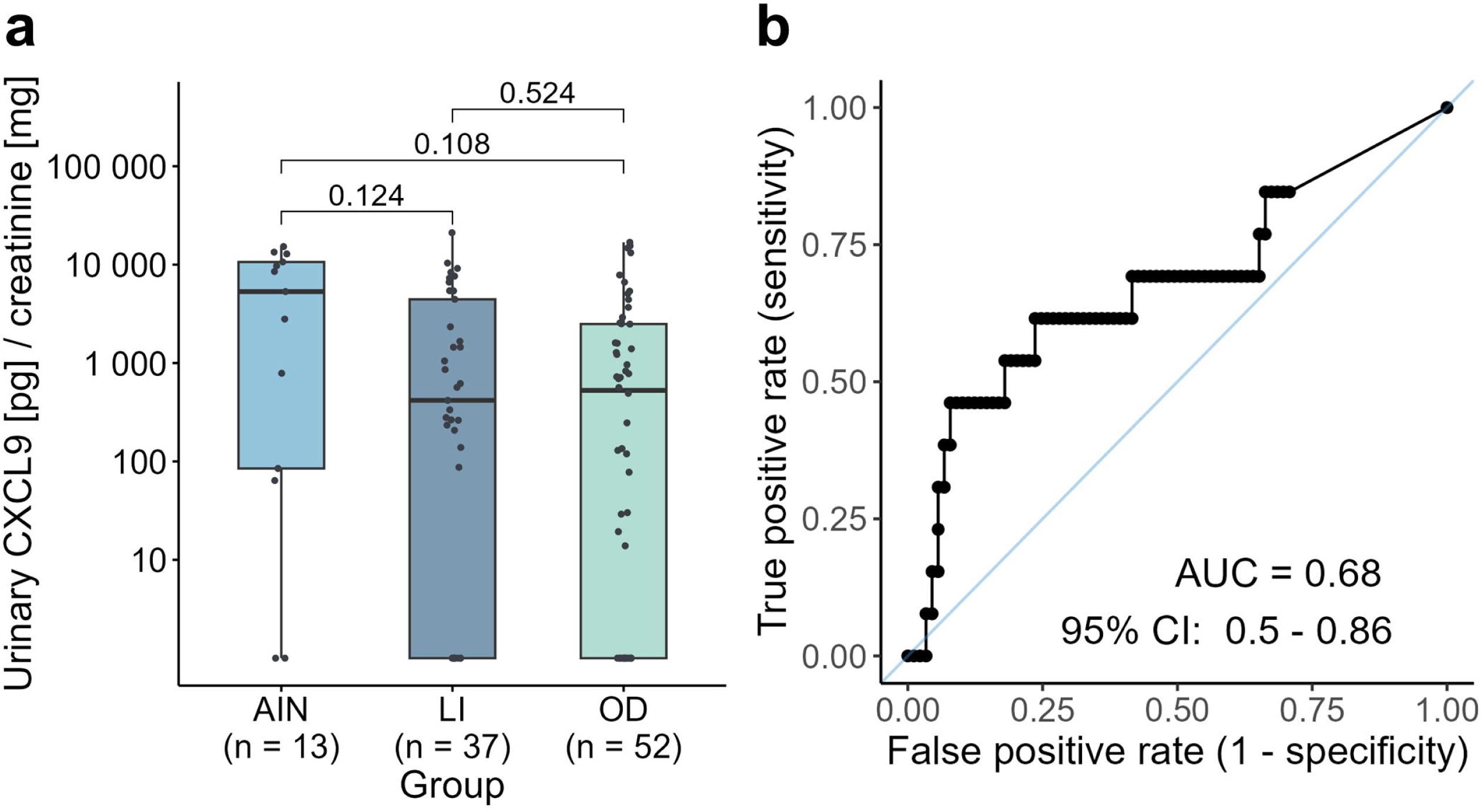
Comparison of urinary CXCL9 levels between diagnosis groups. (a) Comparison of creatinine-normalized urinary CXCL9 concentrations between diagnosis groups of the discovery cohort and internal validation cohort. (b) ROC curve analysis showing the diagnostic performanceof urinary CXCL9 in predicting AIN in both cohorts. AIN, acute interstitial nephritis; LI, lymphocytic interstitial infiltrate in primary glomerular disease; OD, other diseases without interstitial involvement, ROC, receiver operating characteristic; AUC, area under the curve; CI, confidence interval.

### Urinary CD4^+^ EM CD38^+^ HLA-DR^+^ T cell counts correlate with the extent of renal T cell infiltration in AIN

To further examine the relationship between urinary T cells and renal T cell infiltration, immunofluorescence analyses were performed on kidney biopsy specimens from the discovery and the internal validation cohorts. A total of 25 biopsy specimens were stained for DAPI, CD4, and CD38, including 16 specimens from patients with AIN and 9 from patients without AIN (**Figure 4a and b**). In patients with AIN, urinary CD4^+^ EM CD38^+^ HLA-DR^+^ T cell counts showed a positive correlation with the number of CD4^+^ T cells infiltrating the corresponding kidney biopsy (R = 0.63, *P* = 0.009; **Figure 4a**), as well as with biopsy-detected CD4^+^ CD38^+^ T cells (R = 0.68, *P* = 0.004; **Figure 4b**). Representative immunofluorescence images of kidney biopsy tissue stained for DAPI (nuclei, blue), CD4 (green), and CD38 (red) in patients with high-grade or lowDgrade interstitial inflammation are shown in **Figure 4c and d**. Furthermore, there was a positive correlation of urinary and biopsy detected CD4^+^ T cell counts and CD4^+^ CD38^+^ T cell counts (**Supplementary Figure S5c and d**). In contrast, no significant correlations were observed in patients without AIN. Taken together, these findings indicate that urinary CD4^+^ EM CD38^+^ HLA-DR^+^ T cell counts reflect the extent of renal T cell infiltration in AIN.

**Figure 4.**
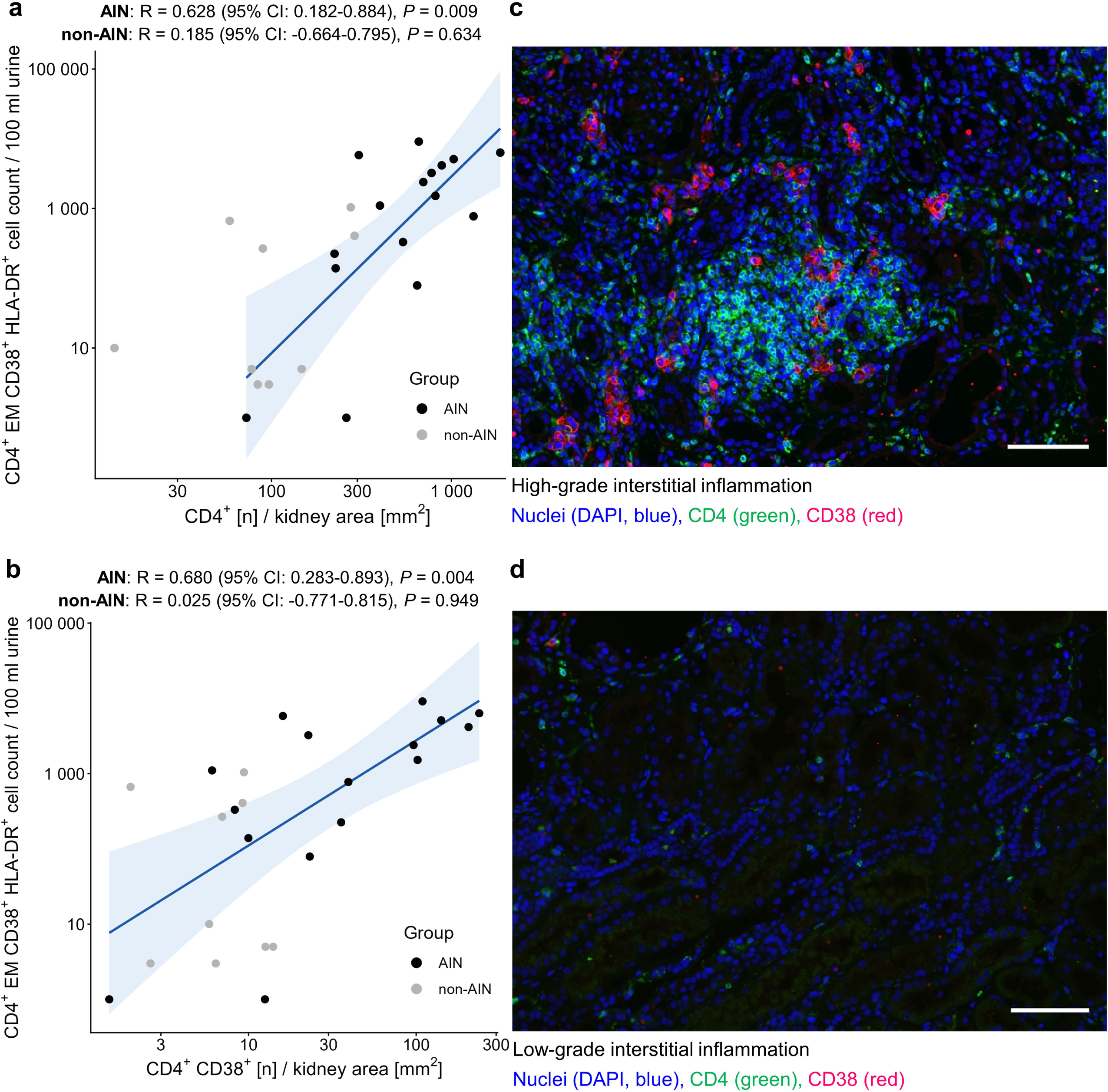
Relationship between urinary T cell counts and renal T cell infiltration. (a-b) Spearman correlation scatter plots between CD4^+^ EM CD38^+^ HLA-DR^+^ T cell counts per 100 mL urine and biopsy-stained total CD4^+^ or CD4^+^ CD38^+^ cells per kidney area in pm^2^, respectively. Blue line represents linear regression with 95% CI. (c-d) Representative immunofluorescence images of kidney biopsy tissue stained for DAPI (nuclei, blue), CD4 (green), and CD38 (red) in patients with high-grade or lowDgrade interstitial inflammation, respectively. White scale bars indicate 100 µm. AIN, acute interstitial nephritis; R, Spearman‘s rank correlation coefficient, *P*, p-value; CI, confidence interval.

### Prior corticosteroid treatment influences urinary CD4^+^ EM CD38^+^ HLA-DR^+^ T cell counts

Although urinary CD4^+^ EM CD38^+^ HLA-DR^+^ T cell counts showed robust diagnostic accuracy for AIN, some AIN patients presented only with minimal numbers of these cells in the urine. We therefore examined the impact of preceding corticosteroid treatment on urinary CD4^+^ EM CD38^+^ HLA-DR^+^ T cell counts. AIN patients who had received corticosteroid treatment for 1 day to 3 weeks before urine sampling exhibited significantly lower urinary CD4^+^ EM CD38^+^ HLA-DR^+^ T cell counts compared to patients without prior corticosteroid treatment (*P* < 0.01; **Figure 5a**). Consistent with this finding, exclusion of patients with prior corticosteroid treatment further improved the diagnostic performance of the biomarker in both the discovery and internal validation cohorts, whereas no change was observed in the external validation cohort, where no patients had received corticosteroids before urine sampling (**Figure 5b-d**). In pooled analyses of all cohorts, the ROC-AUC increased to 0.91 (95% CI, 0.84-0.98; **Figure 5b-d**). Under these conditions, a higher cutoff of 374 activated CD4^+^ EM T cells per 100 mL urine was associated with a sensitivity of 84% and a specificity of 86%, as derived from Youden’s index.

**Figure 5.**
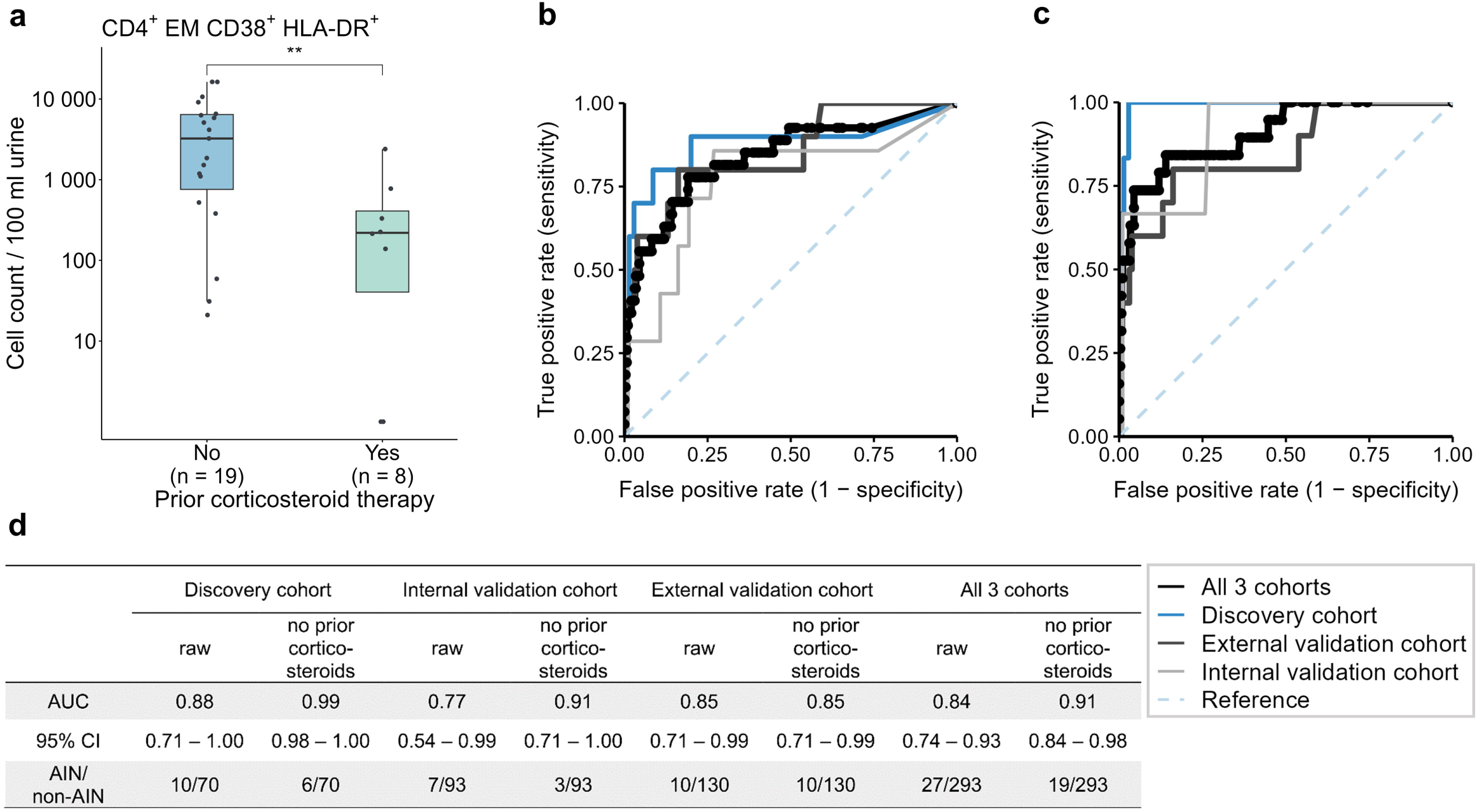
Diagnostic performance of urinary CD4^+^ EM CD38^+^ HLA-DR^+^ T cells for AIN. (a) Counts of urinary CD4^+^ EM CD38^+^ HLA-DR^+^ T cells per 100 mL of urine in dependence on prior corticosteroid therapy in AIN for all AIN patients. (b) ROC curve analysis showing the diagnostic performance of this T cell population in predicting AIN across all cohorts, and (c) after exclusion of AIN patients with prior steroid therapy. (d) Table displaying the corresponding AUC values, accompanied by the 95% CI and numbers of patients with and without AIN, respectively. Significance levels: \*\**P* < 0.01. ROC, receiver operating characteristic; AUC, area under the curve; CI, confidence interval; AIN, acute interstitial nephritis.

### Distribution of urinary CD4^+^ EM CD38^+^ HLA-DR^+^ T cell counts across AIN etiologies and primary kidney diseases

To assess whether the etiology of AIN impacts the amount of urinary CD4^+^ EM CD38^+^ HLA-DR^+^ T cells, AIN patients were stratified by the presumed causative factor. Overall, similar amounts of urinary marker counts were observed across different AIN etiologies. Only the two AIN cases attributed to traditional Chinese medicine exposure tended to display lower urinary CD4^+^ EM CD38^+^ HLA-DR^+^ T cell counts than other causes of AIN (**Figure 6a**).

**Figure 6.**
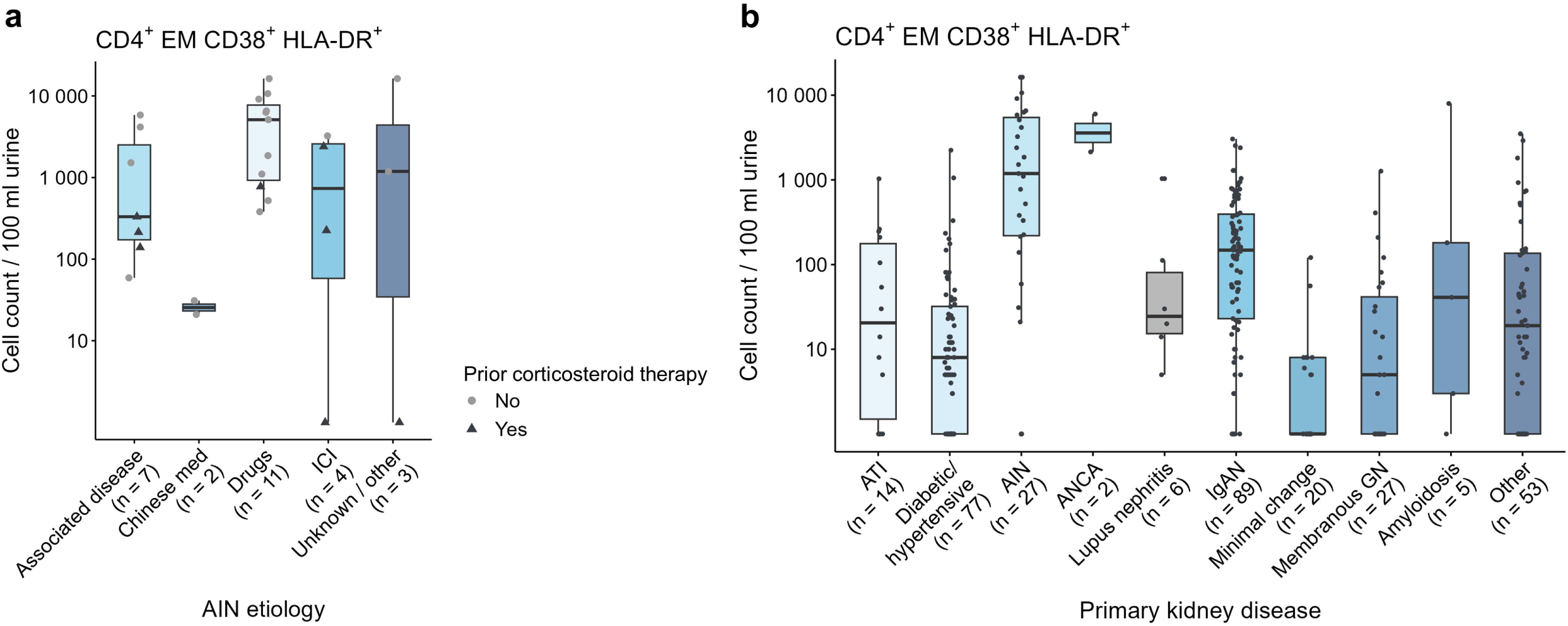
Urinary CD4^+^ EM CD38^+^ HLA-DR^+^ T cell counts per of 100 mL urine by group. (a) AIN etiologies. (b) Primary kidney diseases. Other primary kidney diseases include, but are not limited to Henoch-Schonlein purpura, focal segmental glomerulosclerosis, light chain deposition disease, and obesity-related glomerulopathy. AIN, acute interstitial nephritis, ICI, immune checkpoint inhibitors; ATI, acute tubular injury; ANCA, anti-neutrophil cytoplasmic antibodies-associated glomerulonephritis; IgAN, IgA nephropathy; GN, glomerulonephritis.

Finally, we assessed urinary marker counts across underlying primary kidney diseases. Marker counts partially overlapped between patients with AIN and those with other inflammatory kidney diseases, including IgA nephropathy, lupus nephritis, and ANCA-associated vasculitis (**Figure 6b**), many of whom belonged to the LI group.

## Discussion

AIN is a frequently suspected and potentially treatable cause of AKI. Although most patients respond well to corticosteroid therapy and - where applicable - withdrawal of the causative agent, a favorable outcome critically depends on timely diagnosis and treatment.^2,3^ This study demonstrates that urinary T cells and in particular urinary CD4^+^ EM CD38^+^ HLA-DR^+^ T cells discriminate AIN from kidney diseases without interstitial inflammation or secondary interstitial infiltrates. This robust diagnostic performance outperformed the detection of other immune cells in urine, including monocytes and eosinophils as well as CXCL9 concentrations in urine and was confirmed in two independent validation cohorts. Notably, diagnostic performance further improved after exclusion of patients with prior corticosteroid treatment.

Multiple urinary biomarkers have previously been investigated for their diagnostic utility in AIN. Drug-induced AIN is considered to present an “allergic-like” reaction involving eosinophils, which can be observed in some renal biopsy specimens from patients with AIN.^22^ Accordingly, early studies proposed urinary eosinophils as a diagnostic biomarker for AIN.^23,24^ However, subsequent larger studies demonstrated that the diagnostic performance of urinary eosinophils was unsatisfactory. In a study by Ruffing et al., including 152 patients with pyuria and 51 patients with suspected AIN, the sensitivity of urinary eosinophils for detecting AIN was only 40%, and the positive predictive value did not exceed 38%.^25^ This observation was further supported by a large retrospective cohort study by Muriithi et al., which analyzed 566 patients undergoing kidney biopsy for AKI and demonstrated limited diagnostic utility of urinary eosinophils for AIN, with a sensitivity of only 30.8%, a specificity of 68.2%, and a particularly low positive predictive value of 15.6%.^9^ Consistent with these findings, our study also showed no significant differences in urinary eosinophil counts among the AIN, LI, and OD groups in the discovery cohort.

Given that the histopathological hallmark of AIN is dense lymphocytic interstitial infiltration, T cells are presumed to represent key effector cells in disease pathogenesis. Early immunohistochemical studies demonstrated that the interstitial infiltrates in AIN is predominantly composed of T lymphocytes, including both CD4^+^ and CD8^+^ subsets, as initially suggested by descriptive case analyses and subsequently confirmed in a quantitative immunohistological study across different AIN etiologies.^26,27^ Studies focusing on immune checkpoint inhibitor-associated AIN further supported the central role of T cells by reporting increased circulating frequencies of multiple CD4^+^ T cell subsets, including CD4^+^ effector memory T cells.^28^ Notably, Singh et al. demonstrated the potential of urinary CD4^+^ and CD8^+^ T cells as biomarkers in a small cohort of patients with immune checkpoint inhibitor-associated AIN and, in one patient, reported a closer correspondence of T cell receptor repertoires between kidney biopsy specimens and matched urine samples than between tissue and peripheral blood.^29^ Most recently, a prospective single-center study by Laas et al. demonstrated that urinary CD4^+^ T cells are markedly increased in patients with Sjögren’s disease (SjD) associated tubulointerstitial nephritis and can robustly discriminate SjD-TIN from SjD-related CKD and other control groups.^30^ Importantly, no other urinary leukocyte subset or routine clinical parameter achieved comparable diagnostic discrimination. The authors further reported a pronounced decline in urinary CD4^+^ T cell counts during both early (median 37 days) and longer-term (median 360 days) follow-up after initiation of immunosuppressive therapy,^30^ supporting their potential utility as a biomarker to monitor treatment response. Additionally, a strong correlation between urinary T cell frequencies and the severity of tubulitis in matched kidney biopsy specimens was observed.^30^ Together, these findings support the concept that urinary T cells reflect intrarenal lymphocytic inflammation. In line with these observations, our study demonstrated that activated CD4^+^ EM T cell counts correlated both with the diagnosis of AIN and with the extent of renal T cell infiltration.

Beyond cellular biomarkers, soluble immune mediators have also been extensively investigated as non-invasive diagnostic tools for AIN, based on the assumption that cytokines released by intrarenal immune cells may be detectable in urine and reflect renal inflammation. In this context, Moledina et al. analyzed 12 selected urine and plasma cytokines and identified TNF-α and IL-9 as diagnostic biomarkers of AIN.^11^ In a subsequent proteomic screening of 180 urinary proteins, the same group further validated CXCL9 as a promising biomarker with good diagnostic performance for distinguishing AIN from other kidney diseases.^12^ These findings were subsequently extended to a multicenter study on immune checkpoint inhibitor-associated AIN, where urinary CXCL9 also showed good diagnostic performance.^14^ In contrast, in our cohort, urinary CXCL9 did not show robust discriminatory performance separating AIN from other diseases. One possible explanation may be the composition of our control group, which included a large proportion of patients with inflammatory kidney diseases, such as lupus nephritis, ANCA-vasculitis, and IgA nephropathy. CXCL9 is likely also produced locally in at least the very inflammatory subset of these patients, thereby limiting its ability to identify patients with AIN. Nevertheless, CXCL9 may still be useful for distinguishing inflammatory renal diseases, such as AIN, from non-inflammatory causes of AKI, as previously demonstrated in immune checkpoint inhibitor-associated AIN compared with hemodynamic AKI.^13^

Our study has several strengths. Most importantly, the identified urinary T cell biomarker was validated both internally and in an independent external cohort from another country, supporting its robustness and generalizability across different clinical settings. Second, all urine samples were processed using an experimentally validated preservation and storage protocol,^19^ which was applied consistently across all cohorts to ensure comparability of flow cytometric analyses. Third, the use of cellular biomarkers represents another key strength, as it may minimize confounding by effects of other organs and systemic processes on circulating molecules and more directly reflects intrarenal immune processes. This concept was further supported by corresponding immunofluorescence analyses of kidney biopsy specimens.

Nevertheless, several limitations should be acknowledged. Some correlation analyses were not performed across all cohorts, which may limit the comparability and generalizability of specific findings. In particular, immunofluorescence staining of kidney biopsy specimen and CXCL9 immunoassays were not available for all patients. Moreover, the relatively limited number of AIN patients precluded meaningful subgroup analyses by specific AIN etiologies. As a result, the ability of the identified biomarker to differentiate between etiological subtypes of AIN remains unresolved.

In summary, we identified urinary CD4^+^ EM CD38^+^ HLA-DR^+^ T cells, detected by flow cytometry, as a promising cellular biomarker for the diagnosis of AIN. Diagnostic performance was further improved after accounting for prior corticosteroid exposure. In addition, urinary CD4^+^ EM CD38^+^ HLA-DR^+^ T cell counts correlated with the extent of interstitial lymphocytic infiltration in the kidney, supporting their biological relevance. Together, these findings suggest that urinary activated CD4^+^ EM T cells may serve as a non-invasive adjunct to renal biopsy for the identification of patients with AIN and for guiding early clinical decision making. Future prospective studies with longitudinal sampling will be essential to establish their role in disease monitoring and to further elucidate the molecular mechanisms linking urinary T cells to intrarenal inflammation, thereby paving the way for their clinical translation into precise diagnosis, therapy, and prevention.

## Supporting information

AIN Paper Supplementary Material

## Data Availability

The full protocol, de-identified data, and analysis scripts will be made available upon publication to ensure transparency and reproducibility.

## Abbreviations

AIN: acute interstitial nephritis AKI, acute kidney injury
CKD: chronic kidney disease
CXCL9: CXC-Ligand 9
TNF-α: tumor necrosis factor-alpha IL-9, interleukin-9
ANCA: anti-neutrophil cytoplasmic antibody
CCM: Campus Charité Mitte
CVK: Campus Virchow-Klinikum
FAHZU: The First Affiliated Hospital, Zhejiang University School of Medicine
ROC: receiver operating characteristic
AUC: area under the curve
CI: confidence interval
LI: lymphocyte infiltrate
OD: other diseases
MOPS: 3-(Morpholin-4-yl) propane-1-sulfonic acid
IU: imidazolidinyl urea
FCS: fetal calf serum
DMSO: dimethylsulfoxide
PBE: phosphate-buffered saline/ bovine serum albumin/ ethylenediaminetetraacetic acid (2,5g BSA and 2mL EDTA (0,5 mmol/L) added to 500mL
PBS: resulting in 2mmol/l EDTA in the final PBE solution)
BMI: body mass index
eGFR: estimated glomerular filtration rate
PCR: protein-to-creatinine ratio
ACR: albumin-to-creatinine ratio
sCr: serum creatinine
IQR: interquartile range
SD: standard deviation
H&E: Hematoxylin and Eosin

## Acknowledgements

We thank all participants, participant families, and other participant support persons that have made these studies possible.

We would like to thank Stefan Söllner (FAU) for his excellent technical assistance in performing and evaluating immunofluorescence double staining in kidney biopsies.

## Author contributions

**Conceptualization:** Enghard P*, Jiang H*, Mirkheshti P*, Sha WX*

**Data curation:** Sha WX*, Mirkheshti P*, Feng S, Jiang H*, Enghard P*, Arzig J

**Formal analysis:** Mirkheshti P*, Sha WX*, Skopnik CM, Goerlich N, Klocke J

**Funding acquisition:** Eckardt KU, Enghard P*, Jiang H*, Chen JH

**Investigation:** Sha WX*, Mirkheshti P*, Feng S, Jiang H*, Enghard P*

**Methodology:** Mirkheshti P*, Sha WX*, Feng S, Russ J, Daniel C, Amann K, Skopnik CM

**Project administration:** Jiang H*, Enghard P*

**Resources:** Enghard P*, Jiang H*

**Software:** Sha WX*, Mirkheshti P*, Skopnik CM

**Supervision:** Enghard P*, Jiang H*

**Validation:** Sha WX*, Mirkheshti P*, Feng S

**Visualization:** Mirkheshti P*, Sha WX*

**Writing – original draft:** Sha WX*, Mirkheshti P*

**Writing –review & editing:** Mirkheshti P*, Sha WX*, Jiang H*, Enghard P*, Daniel C, Eckardt KU, Herrmann SM

*These authors contributed equally.

