## Supplementary material for "Urinary CD4^+^ Effector Memory CD38^+^ HLA-DR^+^ T Cells for Diagnosis of Acute Interstitial Nephritis": AIN Paper Supplementary Material

**Supplementary Methods**

### Urine sample collection and processing

Urine samples with a median volume of 60 mL were usually collected within 72 h prior to renal biopsy and transported at 4 °C to local labs. If pre-biopsy samples were unavailable, urine samples were collected no earlier than 24 h and no later than 7 days after kidney biopsy. A total of 8 urine samples were obtained after initiation of corticosteroid therapy for AIN, with treatment duration ranging from 1 day to 3 weeks prior to sample collection. Within 3 h after collection, urine samples were fixed for storage at 4 °C for up to 6 days. For long-term preservation, samples were subsequently stored at -80 °C. Chemical fixation of urinary cells for flow cytometry was conducted as previously described.^1^ Briefly, in-house prepared MOPS buffer (1 mol/L) was mixed with the urine sample at a ratio of 1:3 (v/v), followed by the addition of powdered Imidazolidinyl urea (IU, 20 g/L of total volume; Sigma-Aldrich). To prepare the MOPS buffer, 209.5 g 3-(Morpholin-4-yl) propane-1-sulfonic acid (MOPS; Carl Roth), 8.2 g sodium acetate (Carl Roth), and 18.5 g ethylenediaminetetraacetic acid (EDTA; Sigma-Aldrich) were dissolved in 1000 mL deionized water and adjusted to pH 7.0 with sodium hydroxide (Carl Roth). This preservation method allows a storage at 4 °C for up to 6 days without affecting the following staining. Samples were then centrifuged at 600 × g for 8 min, and the resulting cell pellets were frozen at -80 °C in freezing medium containing 90% FCS and 10% DMSO for a median duration of 6 months. For CXCL9 immunoassays, unfixed urinary supernatants were aliquoted (3 × 1 mL each) and stored at -80 °C for a median duration of 6 months.

### Urinary immune cell staining and flow cytometry

Before flow cytometry staining, frozen urine samples were thawed in 1 mL phosphate-buffered saline containing 0.2% bovine serum albumin and 2 mM ethylenediaminetetraacetic acid (PBE). Thawed samples were filtered through a 30 µm cell strainer (Miltenyi Biotec), centrifuged at 600 × g for 8 min at 4 °C, and resuspended in 400 µL PBE. For the discovery cohort, cell suspensions were allocated to a combined monocyte/eosinophil staining panel and a T-cell staining panel (200 µL per panel, including 50 µL for control stainings and 150 µL for complete stainings). For the internal and external validation cohorts, only the T cell panel was performed (100 µL for control stainings and 300 µL for complete stainings). Human Fc receptor blocking solution (Miltenyi Biotec at Charité Berlin and eBioscience at FAHZU) was then added, and samples were incubated for 20 min at 4 °C prior to monocyte and eosinophil staining and at room temperature prior to T cell staining.

Information of antibodies used for flow cytometry staining is listed in **Supplementary Table S2**. The following antibody dilutions were used: 1:100 for CD45, CD3, CD4, CD45RO, HLA-DR, CD66b and CD14; 1:50 for CD38 and CD62L; 1:200 for CD16; 1:800 for CD36; and 1:20 for Siglec-8. Each complete staining panel was accompanied by a control staining using CD45 and CD3 for the T cell panel and CD45 and CD66b for the monocyte and eosinophil panel. For the discovery cohort and internal validation cohorts, additional white blood cell staining controls were included.

After light-protected antibody incubation for 30 min at room temperature (T cell panel) or 4 °C (monocyte and eosinophil panel), cells were washed with 1 mL PBE by centrifugation (600 × g, 5 min, 4 °C) and resuspended in 100 µL PBE. Flow cytometric analysis was performed using the BD LSRFortessa Cell Analyzer in Berlin and the Cytek® Aurora Analyzer at FAHZU. Data were analyzed using FlowJo Software Version 10.8 BD for Windows. Gating strategies for eosinophils, monocytes, and T cell subsets are shown in **Figure 1A** and **Supplementary Figure S2**. To normalize immune cell counts to 100 mL of urine, the initial urine sample volume, resuspension volume before blocking, volume allocated to each flow cytometry tube, and volume actually measured during flow cytometry were all recorded for each sample.

### Immunofluorescence staining of renal T cells

Immunofluorescence stainings for CD4 and CD38 were performed on biopsy specimens from 16 AIN patients and 9 specimens from LI and OD patients in the discovery and internal validation cohorts. Formalin-fixed human kidney biopsies were dehydrated in series of alcohol and xylene and finally embedded into paraffin followed by cutting into 2 µm sections. After dewaxing in xylene and rehydration, antigen retrieval was performed by cooking for 2.5 min at 120°C in target retrieval solution pH 6 (DAKO Deutschland GmbH, Hamburg, Germany) using a decloaking chamber (BIOCARE medical, Pacheco, CA, USA). The following primary antibodies were diluted in 1% BSA/Tris-buffer pH 7.4 and incubated over night at 4°C: monoclonal rabbit anti-human CD4 antibody (104R-26; Cell Marque, Rocklin, CA, USA); monoclonal mouse anti-human CD38 antibody (MAB24041, Bio-techne GmbH, Wiesbaden-Nordenstadt, Germany). After washing steps, fluorochrome-labeled donkey anti-rabbit IgG Alexa Fluor 555, goat anti mouse IgG Alexa Fluor 647 secondary antibodies (all purchased from Thermo Fisher Scientific GmbH, Dreieich, Germany) were incubated for 30 min at room temperature followed by washing and covering the sections with TrueView mounting kit (Vector laboratories, Newmark, CA, USA) and digitalized using whole slide scanner Axioscan Z1 and Zen software (both from Zeiss GmbH, Oberkochen, Germany). Analysis of CD4 and CD38 positive cells were performed using the Qupath 0.7.0.^2^ In the Qupath software, the wand tool was used to detect and localize the entire tissue area, and the percentage of positively stained cells was calculated using the pixel classifier.

### Validation of urinary CXCL9 as potential AIN biomarker

Levels of CXCL9 were measured in unfixed urine samples from 45 patients in the discovery cohort and 57 patients in the internal validation cohort. Measurements were performed using the MIG (CXCL9) Human ProcartaPlex Simplex Kit (Thermo Fisher Scientific; EPX01A-10285-901) following the manufacturer’s instructions. The only modification was that urine samples were not centrifuged prior to storage at -80°C. To assess whether this modification affected assay performance, paired aliquots from 10 independently collected urine samples were analyzed using the same immunoassay. For each pair, one aliquot was centrifuged prior to storage, whereas the corresponding aliquot was centrifuged immediately before assay execution. No relevant differences in CXCL9 measurements were observed between the two conditions, indicating that omission of centrifugation before freezing did not affect assay results (**Supplementary Figure S3**). Urinary creatinine concentrations were determined using the R&D Systems Kit by bio-techne (KGE005) to normalize CXCL9 levels to urine concentration. Optical densities were acquired using the SpectraMax i3x microplate reader and analyzed with SoftMax Pro software.

**Supplementary Figures and Tables**

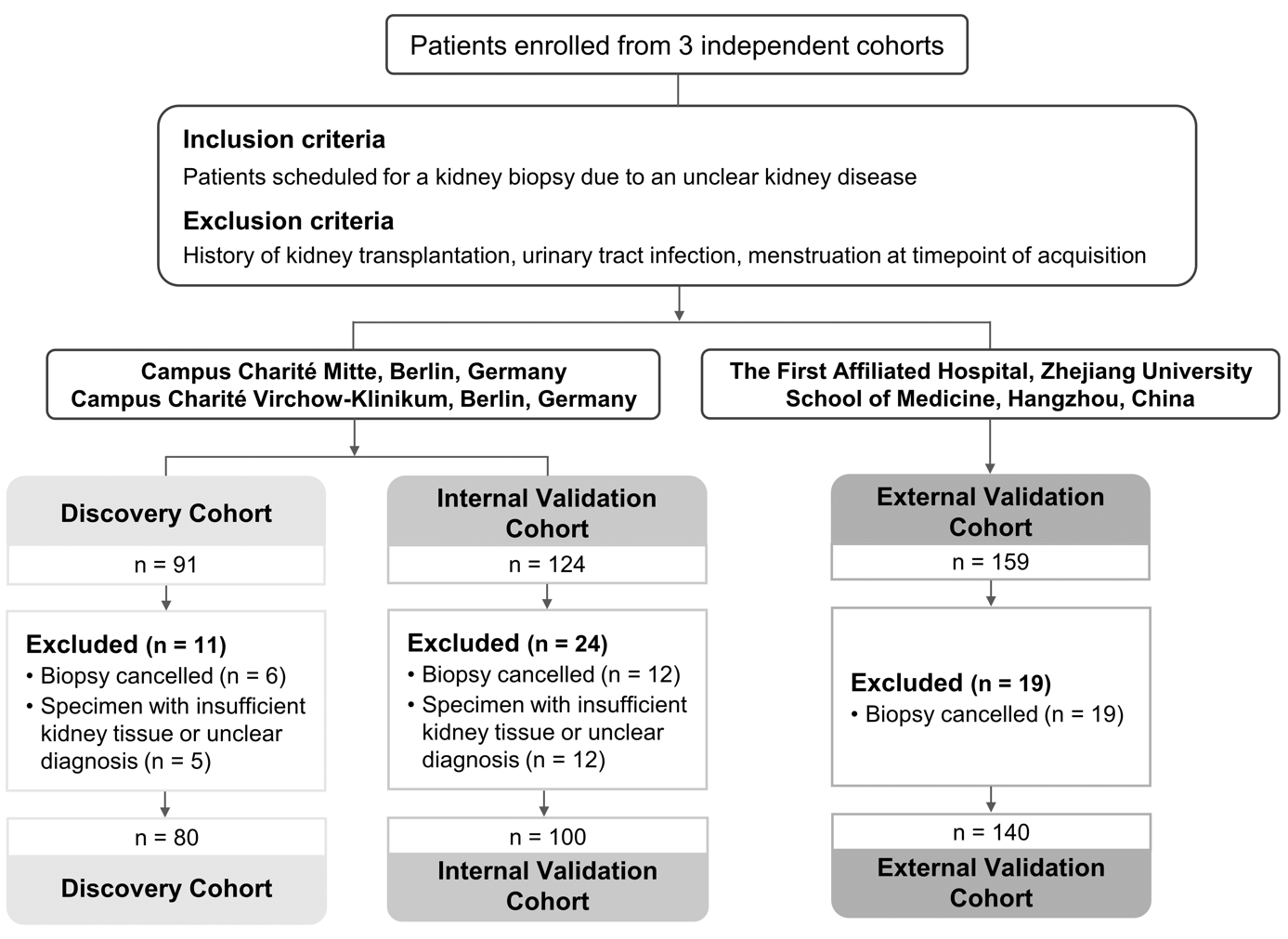

**Supplementary Figure S1. AIN study design**

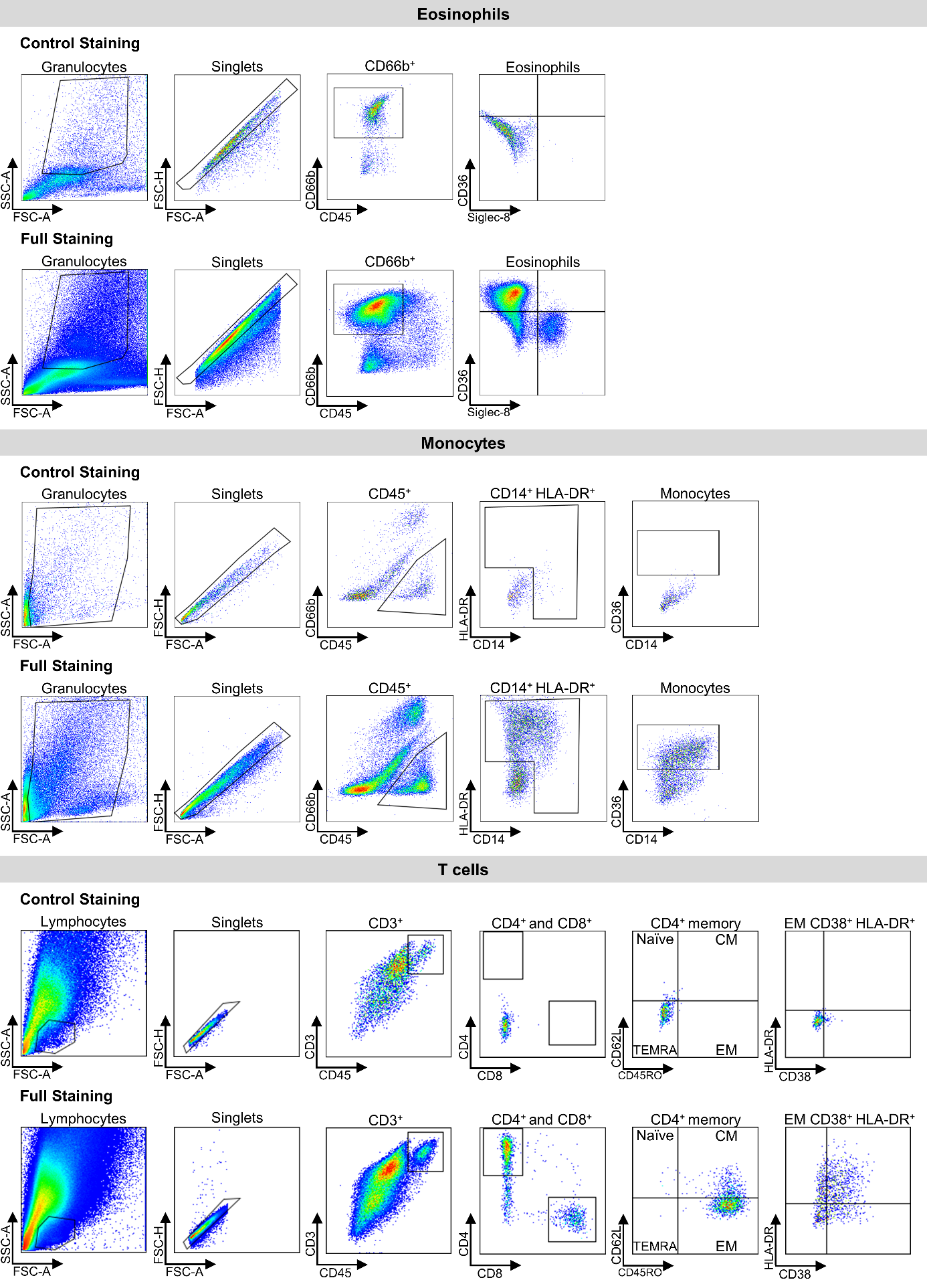

**Supplementary Figure S2. Flow cytometric gating strategies of eosinophils, monocytes, and T cell subsets.** Control stainings were performed for CD45 and CD3. FSC, forward scatter; SSC, sideward scatter; CM, T central memory cells; EM, T effector memory cells; TEMRA, T effector memory cells re-expressing CD45RA.

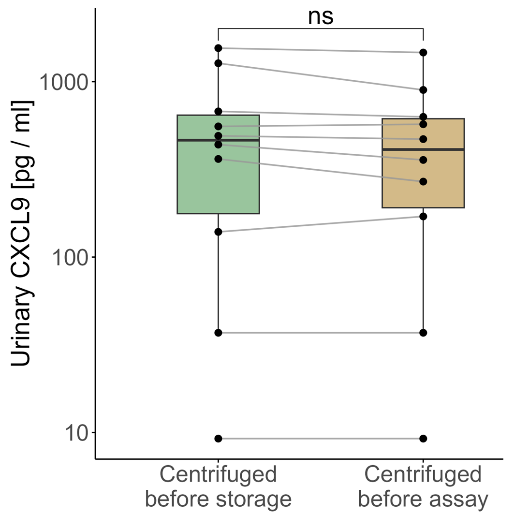

**Supplementary Figure S3. Comparison of urinary CXCL9 levels between diagnosis groups.** Urinary CXCL9 concentrations measured in paired aliquots from the same 10 urine samples to assess whether centrifugation before storage at – 80 °C versus immediately before assay execution affects measured CXCL9 levels.

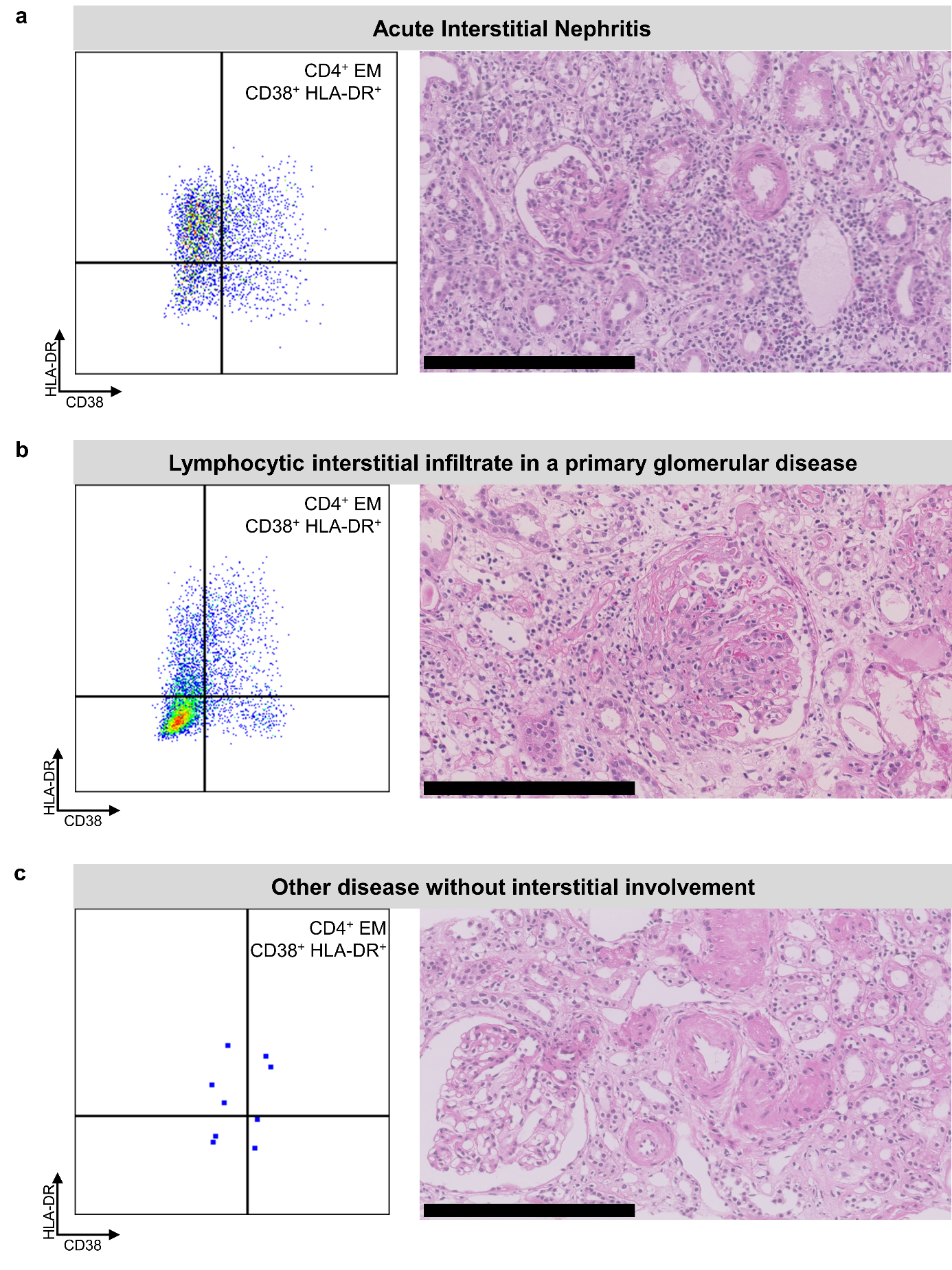
**Supplementary Figure S4. Concordance of renal histology and urinary CD4^+^ EM CD38^+^ HLA-DR^+^ T cells across diagnosis groups.** (a) Representative Hematoxylin- and Eosin-stained kidney biopsy sections from patients with acute interstitial nephritis, (b) lymphocytic interstitial infiltrate in primary glomerular diseases and (c) other diseases without interstitial involvement, together with the corresponding flow cytometric profiles after gating of CD4^+^ EM CD38^+^ HLA-DR^+^ T cells. EM, T effector memory cells. Black scale bars indicate 250 µm.

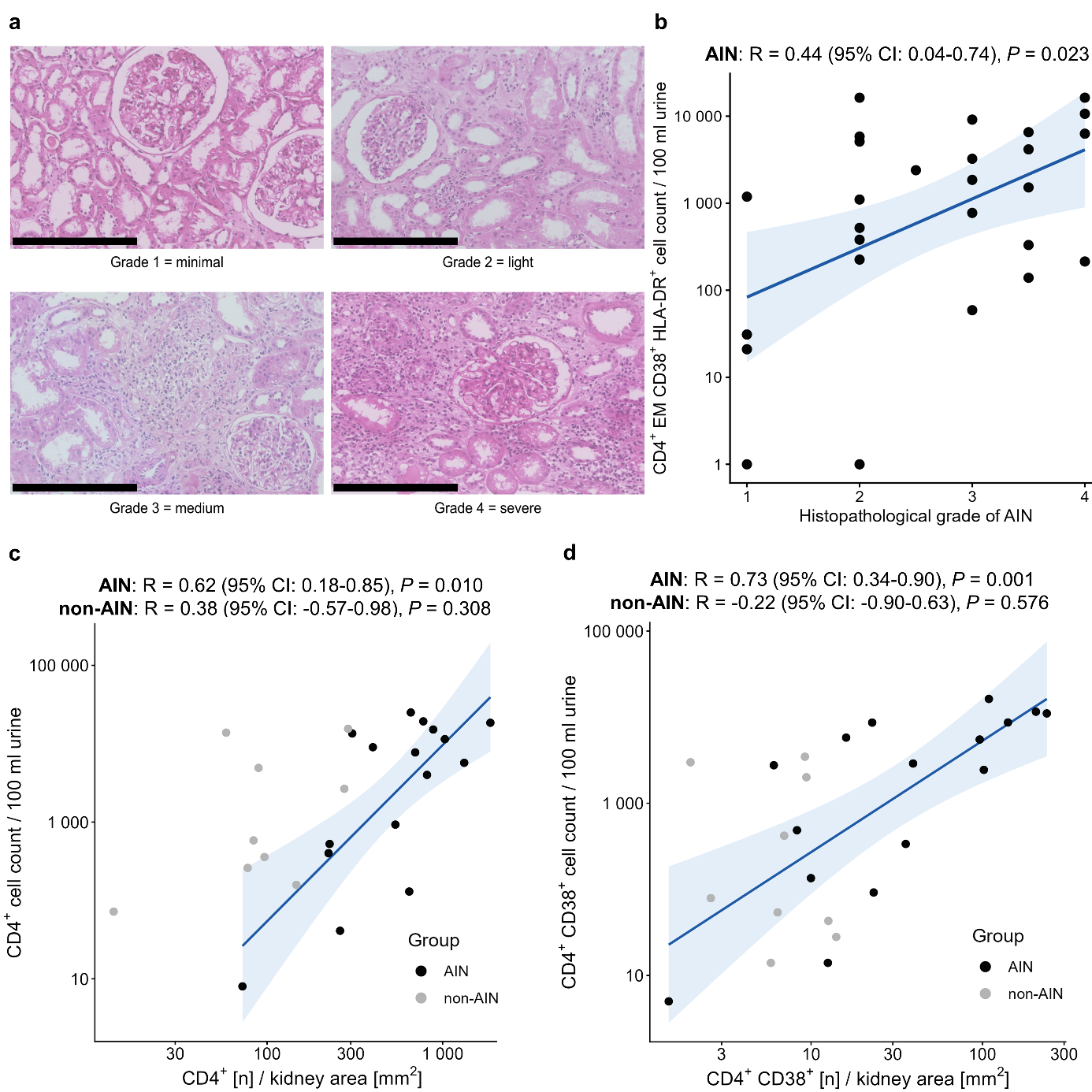

**Supplementary Figure S5. Relationship between urinary T cell counts and renal T cell infiltration.** (a) Representative Hematoxylin- and Eosin-stained kidney biopsy sections from patients with different grades of AIN. Black scale bars indicate 250 µm. (b) Spearman correlation scatter plots between CD4^+^ EM CD38^+^ HLA-DR^+^ T cell counts per 100 mL urine and respective AIN grades. (c) Spearman correlation between CD4^+^ T cell counts per 100 mL urine and biopsy-stained CD4^+^ cells per kidney area in pm^2^. (d) Spearman correlation between CD4^+^ CD38^+^ cell counts per 100 mL urine and biopsy-stained CD4^+^ CD38^+^ cells per kidney area in pm^2^. Blue line represents linear regression with 95% CI. AIN, acute interstitial nephritis; R, Spearman‘s rank correlation coefficient, P, p-value; CI, confidence interval.

**Supplementary Table S1. Distribution of primary kidney diseases among groups**

|  | **AIN** | **LI** | **OD** |
| --- | --- | --- | --- |
| Discovery cohort | AIN | ATI, DN, HN, IgAN, MGN, other^a^ | ATI, DN, HN, IgAN, MGN, MCD, other^a^ |
| Internal validation cohort | AIN | amyloidosis, ANCA, ATI, DN, HN, IgAN, lupus, other^a^ | amyloidosis, ATI, DN, HN, IgAN, lupus, MGN, MCD, other^a^ |
| External validation cohort | AIN | ANCA, IgAN, lupus, MGN, other^a^ | amyloidosis, ATI, DN, HN, IgAN, MGN, MCD, other^a^ |

^a^Including but not limited to Henoch-Schonlein purpura, focal segmental glomerulosclerosis, light chain deposition disease, and obesity-related glomerulopathy.

Abbreviations: AIN, acute interstitial nephritis; ANCA, ANCA-associated glomerulonephritis; DN, diabetic nephropathy; HN, hypertensive nephropathy; IgAN, IgA nephropathy; lupus, lupus nephritis; MCD, minimal change disease; MGN, membranous glomerulonephritis.

**Supplementary Table S2. Antibodies used for urinary immune cell phenotyping^a^.**

**T cell staining panel**

| **Marker** | **Fluorochrome** | **Company** | **Clone** | **Isotype** | **Catalog number** |
| --- | --- | --- | --- | --- | --- |
| CD45 | BUV805 | BD Biosciences | HI30 | mo IgG1, κ | 612891 |
| CD3 | APCeFluor | eBioscience | SK7 | mo IgG1, κ | 47-0036-42 |
| CD4 | PEVio770 | Miltenyi Biotec | REA623 | REA | 130-113-227 |
| CD8 | A647 | BioLegend | SK1 | mo IgG1, κ | 344726 |
| CD38 | A488 | BioLegend | HB-7 | mo IgG1, κ | 356634 |
| HLA-DR | BUV395 | BD Biosciences | G46-6 | mo IgG2a, κ | 564040 |
| CD45RO | PE | BioLegend | UCHL1 | mo IgG2a, κ | 304206 |
| CD62L | BV421 | eBioscience | DREG-65 | mo IgG1, κ | 404-0629-42 |

**Monocyte and eosinophil staining panel**

| **Marker** | **Fluorochrome** | **Company** | **Clone** | **Isotype** | **Catalog number** |
| --- | --- | --- | --- | --- | --- |
| CD45 | BUV805 | BD Biosciences | HI30 | mo IgG1, κ | 612891 |
| CD66b | PEVio770 | Miltenyi Biotec | REA306 | REA | 130-119-768 |
| CD14 | FITC | Miltenyi Biotec | TÜK4 | mo IgG2a, κ | 130-113-146 |
| CD16 | APCVio | Miltenyi Biotec | REA423 | REA | 130-113-390 |
| HLA-DR | BUV395 | BD Biosciences | G46-6 | mo IgG2a, κ | 564040 |
| CD36 | APC | Miltenyi Biotec | AC106 | mo IgG2a, κ | 130-095-475 |
| Siglec-8 | PE | bio-techne | #837535 | mo IgG1 | FAB7975P |

^a^Antibodies included in the multiparameter flow cytometry panel for characterization of urinary T cell subsets, monocytes, and eosinophils are listed together with fluorochromes, companies, clones, isotypes, and catalog numbers.
